# Cancer trends in England in younger adults from 2001-2023: comparing incidence, mortality and stage at diagnosis

**DOI:** 10.64898/2026.06.30.26356042

**Authors:** Amy Berrington de Gonzalez, Esme O’Brien, Zoey Richards, Reuben Frost, Meredith S Shiel, Aislinn Macklin-Doherty, Montserrat Garcia-Closas

**Author notes:** Corresponding Author Sir Richard Doll Building 26 Oakleaf Avenue Sutton SM2 5GP.

## Abstract

**Objectives:** To compare trends in incidence and mortality rates for cancers with rising incidence in younger adults in England, and to assess whether increasing incidence is observed for early-stage, late-stage or both types of disease.

**Methods and analysis:** We used cancer incidence and mortality data from English National Disease Registration Service (2001-2023). Analyses focused on 12 cancers with increasing incidence (on average) in younger adults (20-49 years) and more than 500 cases diagnosed in 2023. Trends were quantified by estimating the average annual percentage changes (AAPCs) and 95% confidence intervals (CI) using Joinpoint regression and by calculating the excess number of cancer cases in 2023 compared to 2001. Age-standardised rates (ASRs) of early (stage 1-2) and late-stage (stage 3-4) disease were compared between 2013 (the earliest year available) to 2023.

**Results:** There were 31,385 cancers diagnosed in younger adults in 2023 compared to 244,384 in older adults. The most common cancers diagnosed in younger adults were female breast (n=8,504), colorectal (n=2,977) and melanoma (n=2,767). Of the 12 cancers that were increasing in younger adults between 2001 and 2023, only two also had increasing mortality rates: endometrial (AAPC[95%CI]= incidence 2.9%[2.4-3.4%] and mortality 4.0%[2.1-6.0%]) and colorectal cancer (AAPC[95%CI]= incidence 3.2%[2.8-3.6%] and mortality 1.9%[1.1-2.7%]). For thyroid cancer mortality rates were stable and for the other cancers (female breast, testicular, ovarian, kidney, brain, prostate, Hodgkin lymphoma and leukaemia) although incidence rates were increasing, mortality rates were decreasing, on average. Of the ten cancers with available stage data six showed increases in incidence rates for both early and late-stage disease between 2013 and 2023. Four cancers showed increases only in late-stage disease (female breast, ovarian, melanoma and Hodgkin lymphoma), while thyroid cancer showed an increase only in early-stage disease.

**Conclusions:** These population-wide analyses of national data from England, combining cancer incidence, mortality and stage-stratified incidence trends, highlight several public health and research priorities. These include identifying the causes of increasing colorectal cancer incidence in younger adults, given the marked increases in mortality and late-stage disease, and of increasing breast cancer incidence, which affects the largest number of younger adults and is increasing only for late-stage disease.

**KEY MESSAGES:** *What is already known on this topic:* Incidence rates for several cancers are rising among younger adults in England. Data from the USA has shown increases in late-stage disease for several cancers increasing in younger adults, but parallel increases in mortality have been observed only for colorectal and endometrial cancer.

*What this study adds:* In younger adults, incidence rates of late-stage disease increased from 2013-2023 for all cancers with available stage data except thyroid cancer. However, only endometrial and colorectal cancer showed parallel increases in mortality rates from 2001-2023. Despite these increases, the absolute numbers of diagnoses and deaths remained relatively small compared to those in older adults (age 50+).

*How this study might affect research, practice or policy:* This study identifies current priorities for research and cancer prevention, interception and early detection in younger adults, underscoring the need for in-depth investigation of cancers with concurrent increases in incidence, mortality, and both for both early- and late-stage disease.

## INTRODUCTION

Cancer incidence rates are increasing for a number of cancers in both younger and older adults in the UK^1^. Several recent reports from the US have compared trends in cancer mortality with incidence and found that mortality rates are increasing in parallel to incidence for some cancers (e.g. colorectal and endometrial cancer), but not for others^2, 3^. Analyses of cancer incidence trends by stage also suggest that some of these increases in younger adults are occurring across all stages of disease (eg breast cancer), but some cancers have increases primarily in earlier stage (eg pancreatic cancer)^4^. Increases in cancer incidence in younger adults that are driven by early-stage disease and without parallel increases in mortality could suggest that increased surveillance or changes in screening guidelines or disease coding could be part of the explanation for the increasing rates^5, 6^.

We previously evaluated trends in cancer incidence in younger and older adults in England and reported that many cancers were increasing in younger adults between 2001 and 2019^1^. We noted, however, that most of these cancers were also increasing in older adults and at a similar rate. Other UK studies have described trends in both cancer incidence and mortality for people aged 35-69 year from 1993-2018^7^, but did not evaluate differences by age group^8^. Here, we examine mortality trends for cancers that are increasing in younger adults and assess whether the increases in incidence are driven by early- or late-stage disease. In addition, we compare trends in younger and older adults to assess whether any of these changes are specific to younger adults. Finally, we report the absolute number of cancer diagnoses and deaths in younger and older adults and the change over time to help inform public health and research priorities.

## METHODS

We used cancer incidence and mortality data from the National Disease Registration Service (NDRS)^9^ from 2001-2023, but excluded 2020 due to evidence of under-reporting/detection of cancer diagnoses that year due to the COVID-19 pandemic^10, 11^.

We first evaluated the total burden of cancer incidence and mortality in younger (age 20-49) and older (age 50-79) adults in 2023 in England for all malignant cancers (except non-melanoma skin cancer, NMSC). We then examined trends since 2001 among cancers with more than 500 cases diagnosed in 2023 in younger adults to identify those increasing only in younger adults. This identified 12 cancers for further analyses: female breast, colorectal, melanoma, thyroid, kidney, leukaemia, oral, brain, Hodgkin lymphoma, ovary, endometrial and prostate cancer, using ICD codes defined by the National Disease Registration Service (NDRS) (Appendix Table 1).

Cancer stage at diagnosis was available annually from 2013-2023 for ten of the twelve cancers (no data for brain tumours or leukemia). However, patterns in missing data varied across the period 2015-2020, partly due to COVID-19 pandemic^10^, creating artefacts in stage trends. We therefore compared rates by stage only between two time points in 2013 and 2023. We classified disease as early-stage (1-2), late-stage (3-4) and unstageable/missing.

### Statistical methods

We evaluated cancer incidence and mortality in younger adults (age 20-49 years) and older adults (age 50-79 years), and by 10-year age-groups. Rates were age-standardised to the European population in 2000.

We used Joinpoint regression for analysis of trends, allowing for up to 4 joins and estimated the average annual percent change (AAPC)^12^. We reported the most recent annual percent change (APC) as well as showing the full trajectory. As trends were generally similar for colon and rectal cancer we report the combined findings for colorectal cancer, but include the separate results in the appendix (Appendix Figure 1).

**Figure 1:**
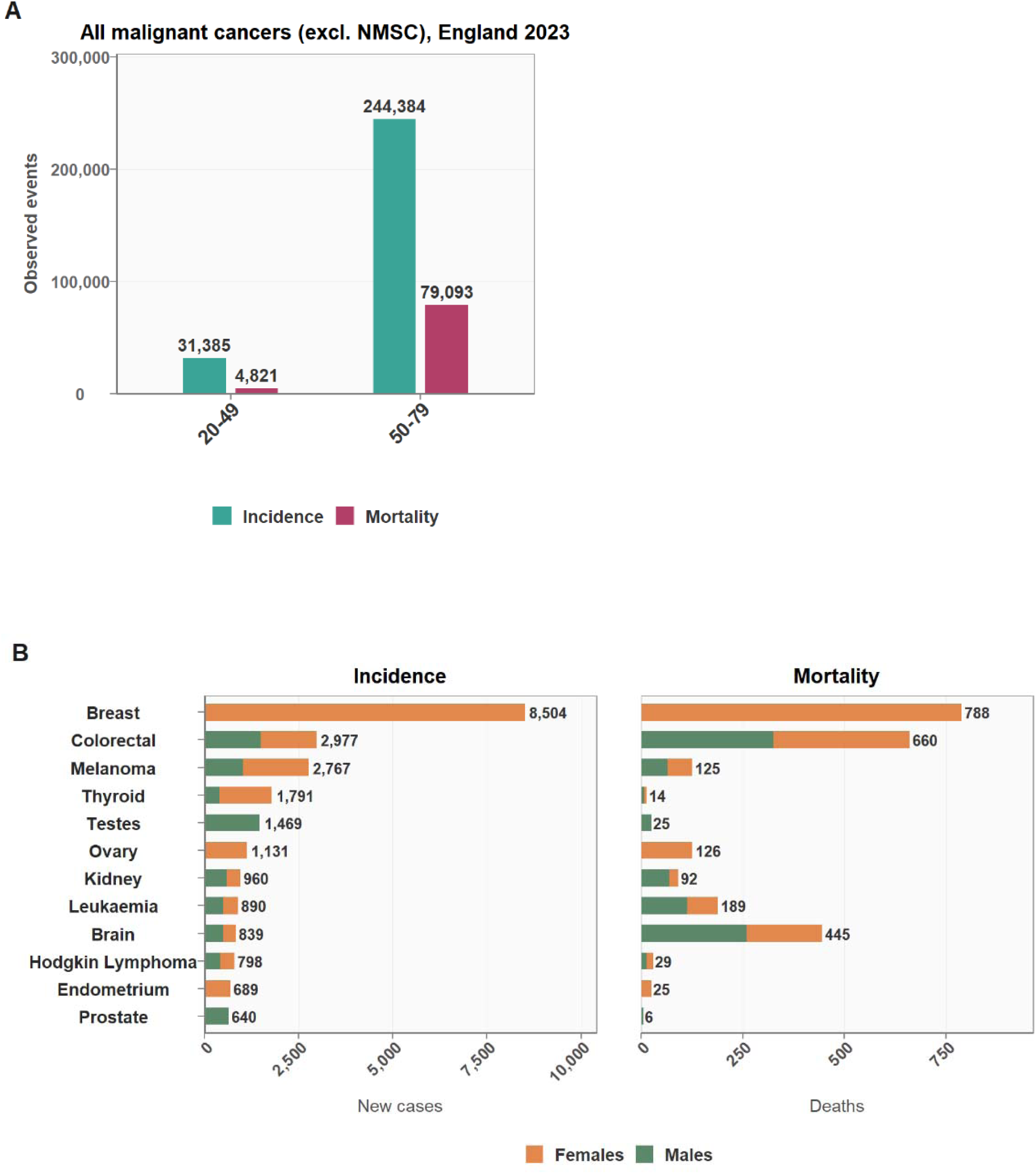
Cancer diagnoses and deaths in England in 2023 for **(A)** all malignant cancers (except non-melanoma skin cancer, NMSC) by age at diagnosis **and (B)** for 12 cancers that increased in 20-49 year olds between 2001 to 2023 by sex

Given the high proportion of unstageable/missing data for stage some cancers (up to 55%) and changes in missing data over time, we conducted two sensitivity analyses assigning all missing stage data to early- and then to late-stage disease.

We estimated the excess cancer cases in 2023 compared to 2001 by applying the age-standardised rates from 2001 to the population structure in 2023.

## RESULTS

In 2023 in England there were 31,385 malignant cancers (excluding NMSC) diagnosed in adults age 20-49 years compared to 244,384 diagnosed in adults age 50-79 years (Figure 1). Approximately two thirds of the diagnoses in younger adults were in females, primarily due to the large number of breast cancers (n=8,504). The second most common cancer in younger adults was colorectal (n=2,977) followed by melanoma (n=2,767), thyroid cancer (n=1,791) and testicular cancer (n=1,469).

There were 6,034 more cancers diagnosed in younger adults in 2023 than expected based on the rates in 2001, and 64% of these additional cases were in females. The largest absolute increases were in colorectal (n=1,604) and thyroid cancer (n=1,228) (Figure 2).

**Figure 2:**
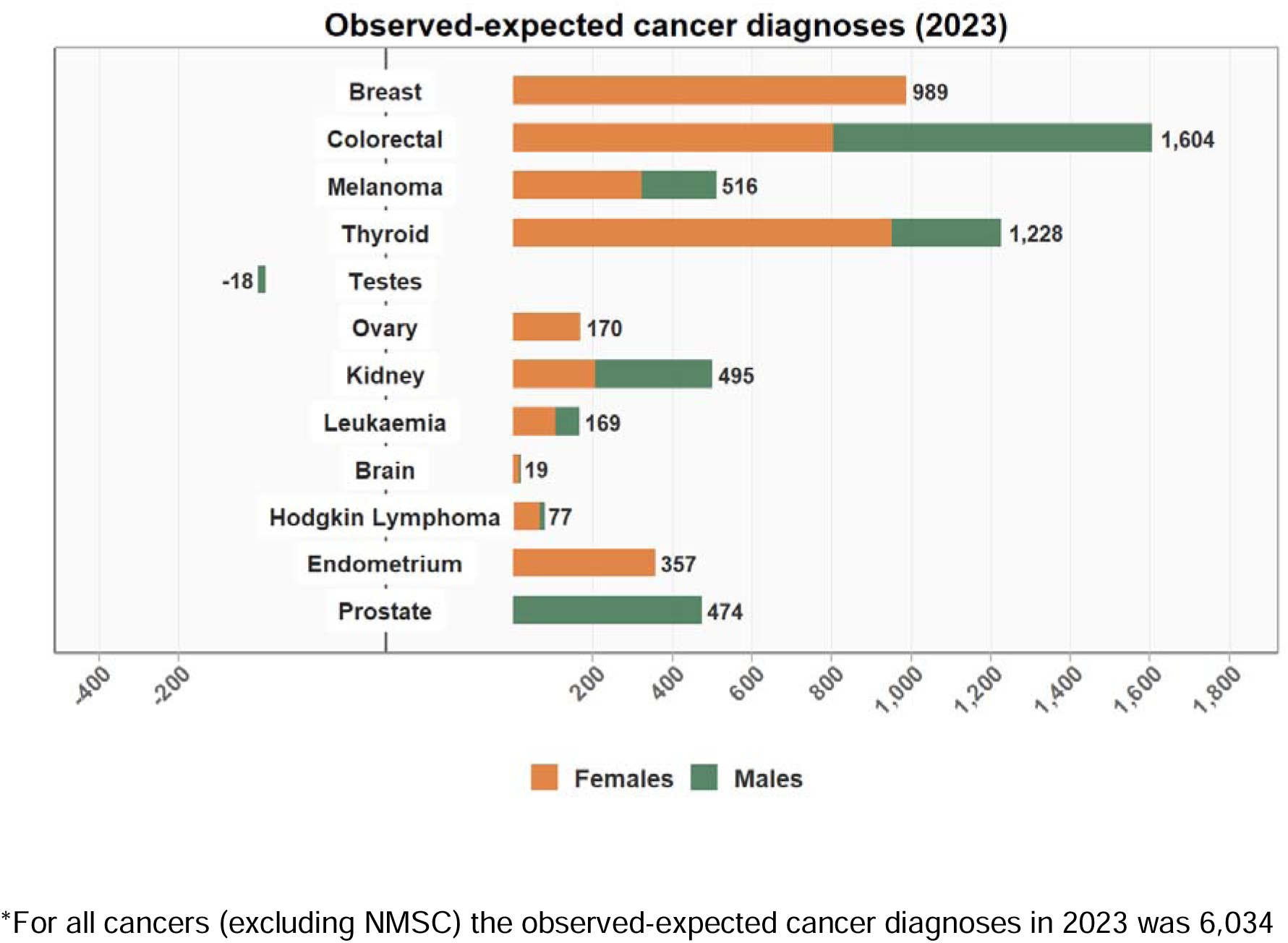
Excess (observed minus expected) cancer diagnoses in England in 2023 by sex, for 12 cancers that increased in 20-49 year olds between 2001 to 2023*. Expected number of cases estimated by multiplying the age-specific cancer incidence rates in 2001 by population counts for 2023.

In 2023 there were 4,821 cancer deaths in younger adults compared to 79,093 in older adults in England. Female breast cancer was the leading cause of cancer death in younger adults (n=788) followed by colorectal (n=660) and brain cancer (n=445) (Figure 1).

When comparing trends in cancer incidence and mortality from 2001-2023 for these 12 cancers that were increasing in younger adults, we observed three distinct patterns (Figure 3). Two cancers had increasing rates of both incidence and mortality in younger adults: endometrial (AAPC[95%CI]= incidence 2.9%[2.4-3.4%] and mortality 4.0%[2.1-6.0%]) and colorectal cancer (AAPC[95%CI]= incidence 3.2%[2.8-3.6%] and mortality 1.9%[1.1-2.7%]) (Figures 3, 4a, Appendix Table 2). For older adults, endometrial cancer incidence and mortality were also increasing, whereas for colorectal cancer incidence and mortality were decreasing.

**Figure 3:**
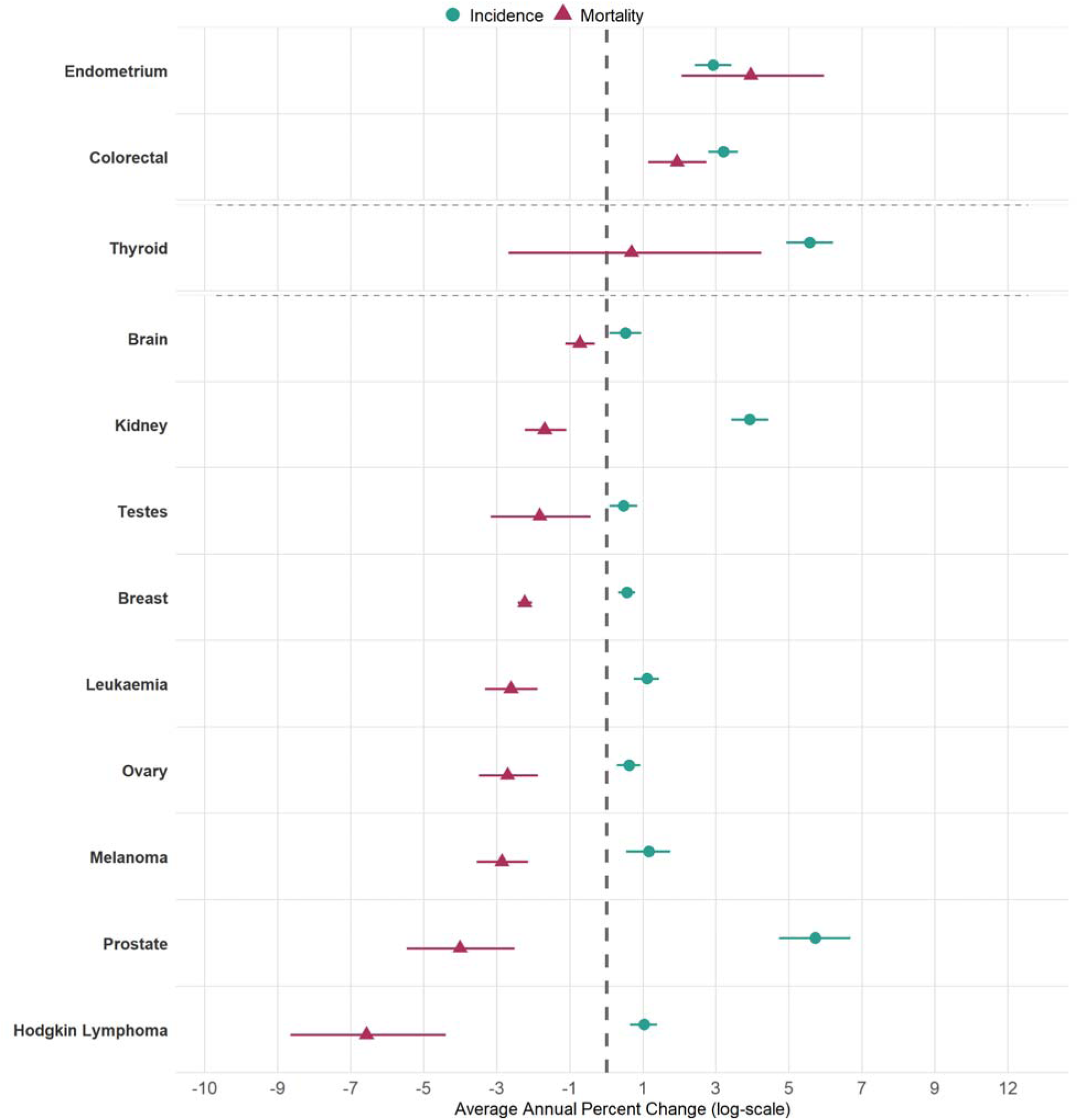
Average annual percent change (AAPC) and 95% confidence intervals in cancer incidence and mortality for 12 cancers that increased in 20-49 year olds between 2001 to 2023. Cancers are ordered by AAPC for mortality rates. Dotted lines classify cancers into three distinct patterns: cancers with increasing incidence and mortality, increasing incidence with constant mortality, and increasing incidence with decreasing mortality.

Thyroid cancer had increasing incidence rates in younger adults but no clear trend for mortality (Figures 3, 4b). In contrast, the other 9 cancers had increasing incidence rates but significantly decreasing mortality rates in younger adults (Figure1, 2c). This included cancers that are traditionally associated with a younger age at onset (testicular, thyroid and brain cancer, leukaemia and Hodgkin’s lymphoma) and those that are traditionally associated with later age at onset (prostate, ovary and kidney cancer). The patterns were similar in older adults for the direction of incidence and mortality trends for all these cancers, except for ovarian cancer that had decreasing incidence as well as mortality in older adults.

Although these 12 cancers had increasing incidence rates overall from 2001-2023, as assessed by the AAPC, only six of them were still increasing significantly in the more recent years in younger adults (as assessed by Joinpoint); these were colorectal, thyroid, endometrial, leukaemia, ovarian and kidney cancer (Figure 4, Appendix Figure 2). For the other six cancers four had decreasing trends in more recent years (breast, melanoma, brain and testes). Analysis of trends by 10-year age-groups were largely consistent with those observed when using the broader age group. Some exceptions included the recent increases in colorectal cancer incidence and mortality, which were primarily observed in 40-49 year olds, recent increases in brain cancer incidence being only in 30-39 year olds and recent significant decreases in melanoma incidence in 20-29 year olds (Appendix Figure 3).

**Figure 4:**
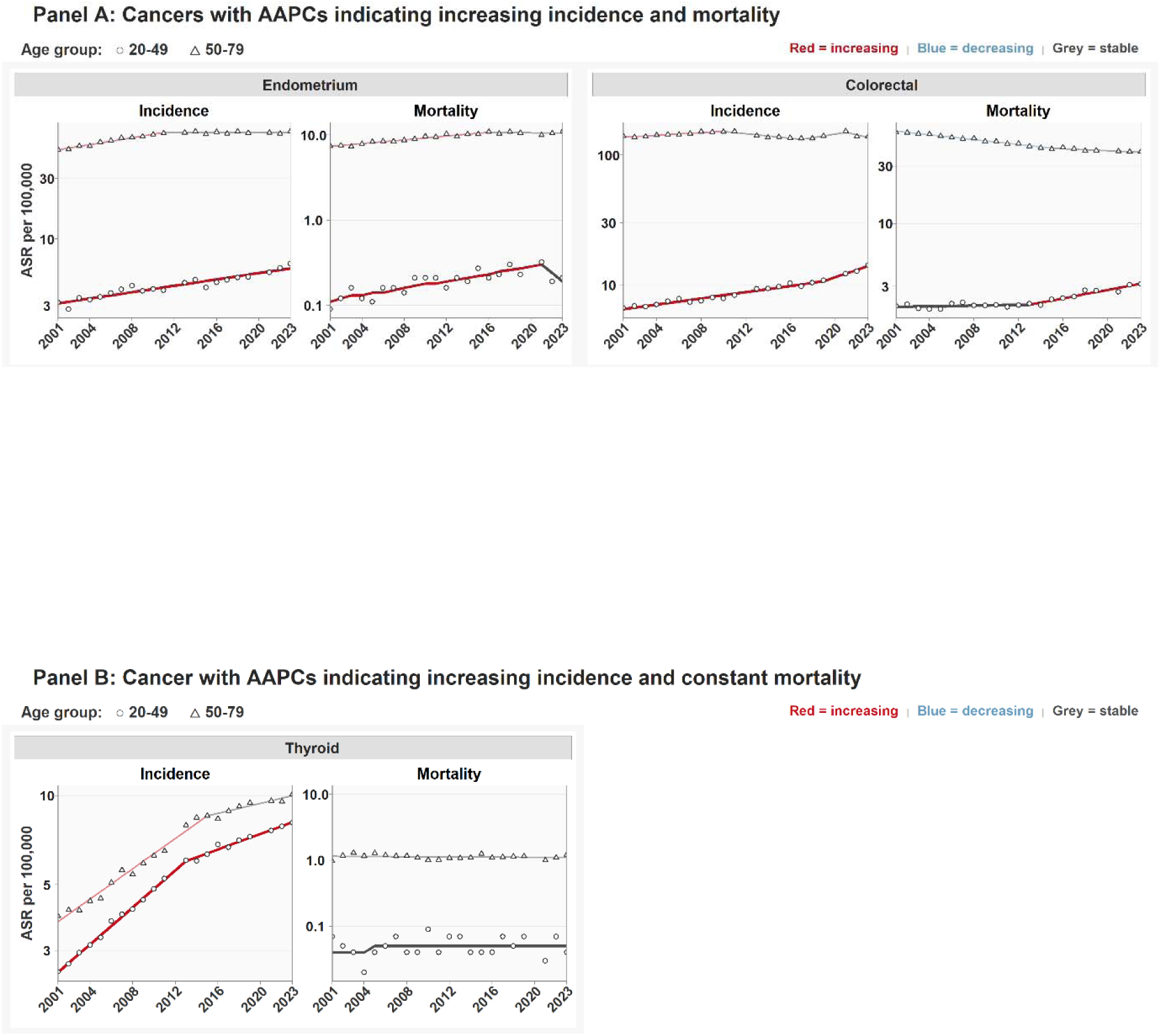
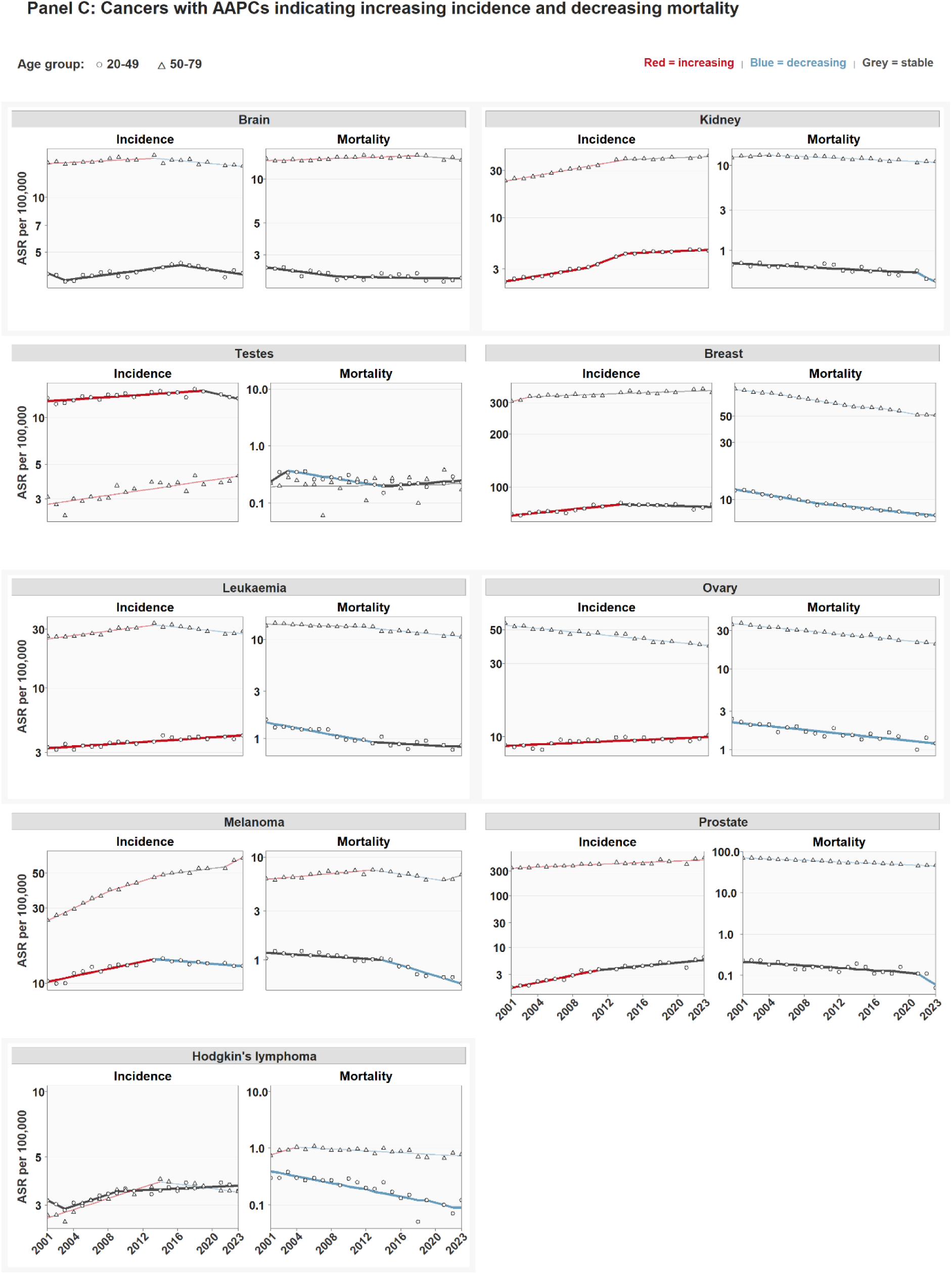
Trends in age-standardised rates (ASR) for cancer incidence from 2001-2023 for 12 cancers that increased in 20-49 year olds between 2001 to 2023 by age group and cancer type in England for **(A)** cancers with AAPCs indicating increasing incidence and mortality, **(B)** cancers with AAPCs indicating increasing incidence and constant mortality, and **(C)** cancers with AAPCs indicating increasing incidence and decreasing mortality. Points represent observed data, and lines represent modeled trends.

We examined the changes in incidence rates by stage at diagnosis from 2013 to 2023 for 10 cancers with stage data, within the three sub-groups with distinct trends patterns in incidence and mortality. For colorectal and endometrial cancers showing increases in both incidence and mortality in younger adults, there were increases in rates of both early- and late-stage disease (Figure 5a). For thyroid cancer showing only increasing incidence, the increase was only in early-stage disease (Figure 5b). For the other seven cancers that had increasing incidence but decreasing mortality rates, three had increases in rates of both early and late-stage disease (kidney, testes, prostate), while four cancers were only increasing in late-stage disease (breast, ovarian, melanoma and Hodgkin’s lymphoma; Figure 5c). Rates of late-stage disease were very low (<1 per 100,000) in younger adults except for breast cancer (4.4 per 100,000) and colorectal cancer (2.7 per 100,000) (Appendix Table 3). However, these are likely underestimates due to the % of missing data.

**Figure 5:**
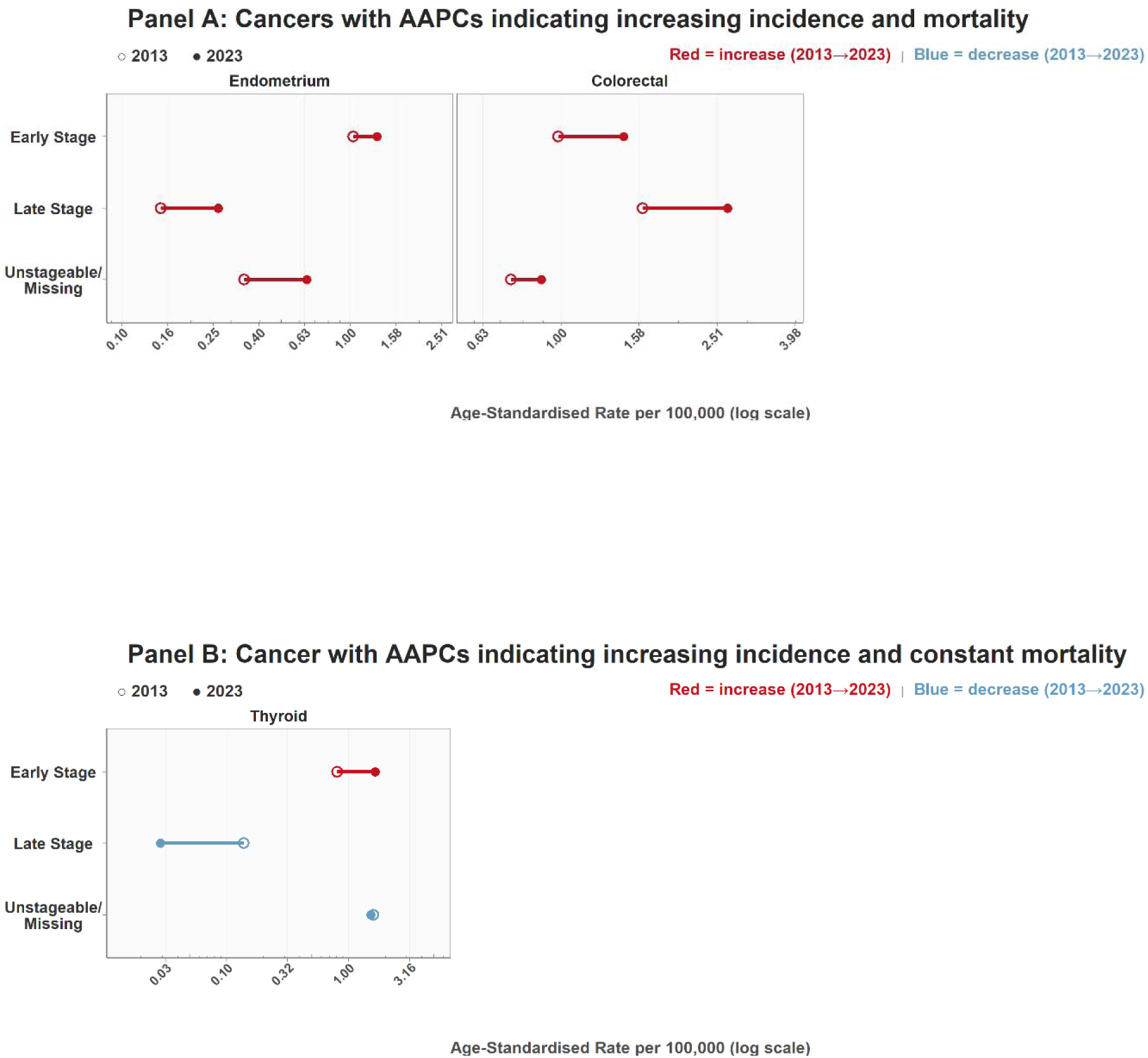
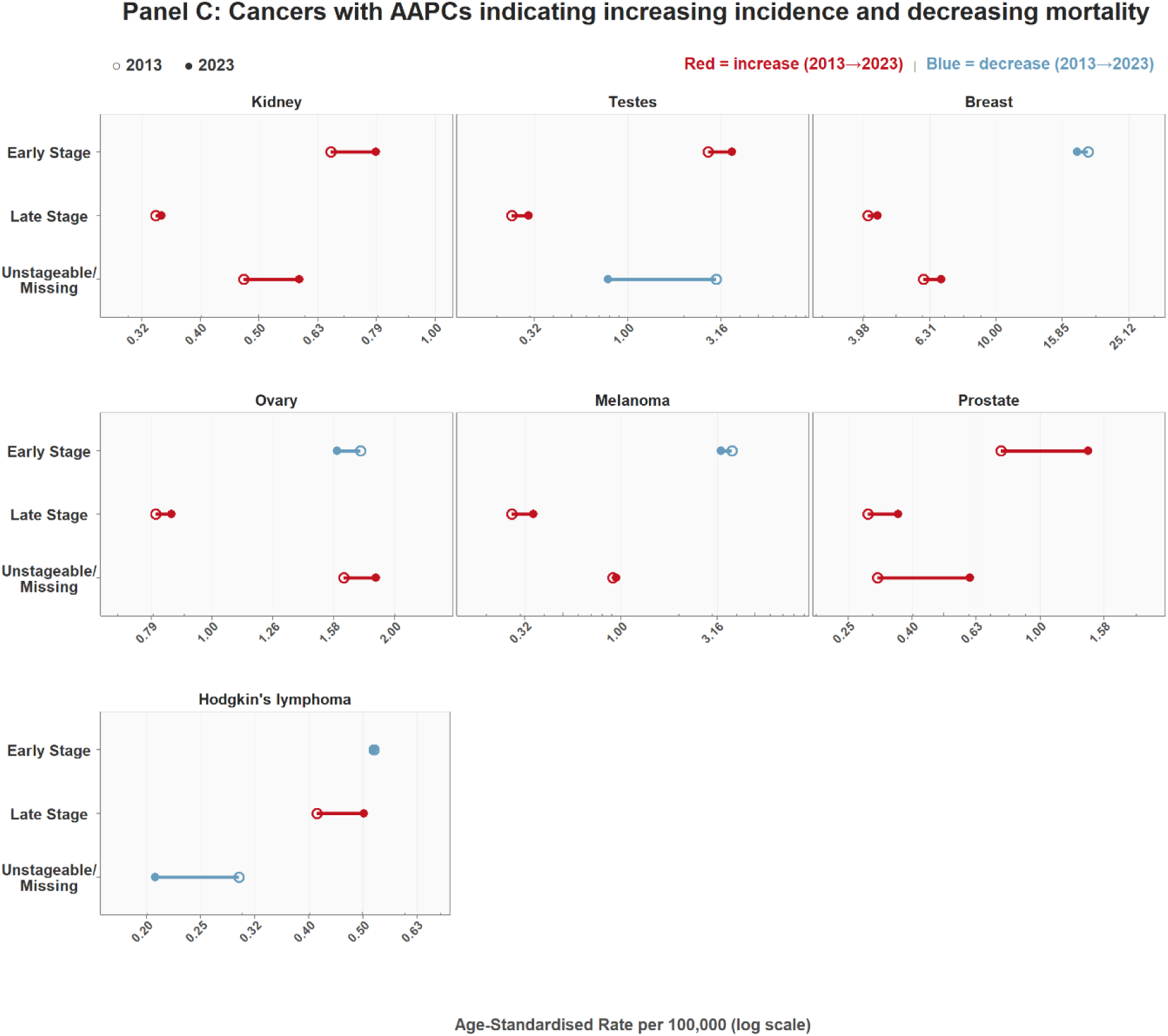
Changes in age-standardised rates (ASR) for cancer incidence in 20-49 year olds between 2013 and 2023 for 12 cancers that increased in 20-49 year olds between 2001 to 2023 by stage at diagnosis for **(A)** cancers with AAPCs indicating increasing incidence and mortality, **(B)** cancers with AAPCs indicating increasing incidence and constant mortality, and **(C)** cancers with AAPCs indicating increasing incidence and decreasing mortality

In 2013 the percentage of cancers diagnosed in younger adults with unstageable/missing stage varied from 55% for thyroid cancer to 15% for endometrial cancer (Appendix Table 4). In 2023 the % of cancers with missing stage was similar or slightly lower, with larger decreases for thyroid cancer (although still 40% missing stage) and testicular cancer (43% in 2013 reduced to 13% missing stage in 2023). In the two sensitivity analyses (reclassifying missing sage as early or late stage) the direction of changes in stage-specific rates were similar. The exception was for ovary and testicular cancer where the reclassification of missing stage data to either early or late stage changed the observed direction of the trend (Appendix Figure 4).

## DISCUSSION

Cancers that are increasing in younger adults in England include those that are traditionally diagnosed at younger ages (eg leukaemia and Hodgkin lymphoma), as well as those that are traditionally later-onset cancers (eg colorectal and endometrial cancer). Only two cancers, endometrial and colorectal cancer, had both increasing incidence and mortality rates in younger adults from 2001 to 2023. Between 2013 and 2023 rates of these cancers increased for both early and late-stage disease. For most other cancers that had increasing incidence rates in younger adults the mortality rates were stable or decreasing on average over the study period. However, all cancers with available stage data had increasing rates of late-stage disease in younger adults except for thyroid cancer.

Patel and Shiels also showed that in the USA most cancers that were increasing in younger adults increased both for early and late stage disease, without parallel increases in mortality^4, 13^. As in our study in England, the exceptions were colorectal and endometrial cancer that had increasing mortality rates in younger adults. This suggests that the increases in incidence for these cancers are unlikely to be explained by increases in surveillance and early detection. It also suggests that advances in treatment have not compensated - or not yet compensated - for the increases in incidence in colorectal and endometrial cancer. For the other cancers with decreasing mortality despite increasing incidence this could be partly due to advances in treatment, and/or that insufficient time has passed to observe increases in mortality. Other population-based studies in the UK have found decreasing mortality for the majority of cancers with increasing incidence in adults^7, 14^, with the exceptions of liver, oral and endometrium cancers in people aged 35-69 from 1993-2018^7^, and liver, endometrium, melanoma and kidney cancer in all adults from 1980-2013^14^. However, age-specific analyses were not presented in previous studies.

To our knowledge, this is the first analysis of trends in cancer rates by stage in younger adults in England, which is important to evaluate the potential role of increased surveillance and early detection. The pattern for thyroid cancer in younger adults in England, with increasing incidence rates limited to early onset cancers and stable mortality rates, is consistent with overdiagnosis due to increased surveillance^14–16^. The colorectal cancer screening programme in England started in 2012 for people age 60+ and was expanded to age 50+ in ^17^. High-risk screening for those with a family history was introduced in 2020 for individuals aged 40+ and this could explain some of the recent increased incidence rates for the 40-49 year age-group. Previous analyses of the long-term trends in colorectal cancer incidence in England (1974-2015) in younger adults found that the increases started in the 1990s with increases driven by distal cancers in the youngest adults but ^18^. There have been some changes in breast screening in England that might have increased breast cancer rates in younger women during the study period. The very high-risk screening program was formalised in England 2013 and the number of women screened has increased from about 1,000 in 2013 to 10,000 in 2023, but the 200 cancers diagnosed in this population in 2023 is a very small fraction of the ^19^. In addition, the AgeX trial randomised women in England to receive breast screening earlier (age 47) ^20^. However, there was no increase in breast cancer incidence in 40-49 year olds during this period (Appendix Figure 3c). As the increasing breast cancer rates in younger adults were primarily for late-stage disease this suggests that increased screening is unlikely to be the explanation and further research into the causes is warranted.

Kidney, testes and especially prostate cancer had increases in early-stage disease consistent with increases in surveillance- e.g. incidental findings from imaging in kidney and testes, and PSA testing for prostate cancer^5, 13^. The increasing incidence of early-onset kidney cancer in England was reported previously to have increased more than double since the 1980s^21^. However, we found that increases were not restricted to early-stage disease. In fact, incidence for late-stage disease in younger adults increased for all cancers studied except thyroid cancer, and the increases were restricted to late-stage disease for several cancers (breast, ovarian cancer, Hodgkin lymphoma and melanoma). Mortality rates were decreasing for most of these cancers, likely due to improvements in treatment (e.g. immunotherapy for multiple cancers and PARP inhibitors for breast and ovarian cancers^22, 23^), consistent with findings in adults aged 35-69^7^. However, the increases in late-stage disease are concerning and could slow or reverse the favourable mortality trends, warranting further evaluation.

The increasing incidence for some cancers could partly reflect reclassification and improved recognition of neuroendocrine tumours (NETs) in young people, particularly for colon, rectum and appendiceal cancers, which have the highest NET incidence^24^. This is due to changes in WHO classification and cancer registry coding since the 2000s that have led to NETs been increasingly classified as^25–27^. Of note, a recent study finding that NETs represent 4-20% of all colorectal cases and 8-34% of rectal cancer cases in^28^. Histological subtype was not available in the publicly available NDRS data we used for the current analysis and future research should evaluate the impact of these changes on the incidence rates.

The strengths of this analysis include national cancer registry and mortality data with high-levels of completeness during the study ^29^. Analysis of changes in the cancer incidence rates by stage also provided additional insights into the potential ^5, 13, 16^, whilst avoiding the limitations of previous analyses that have ^30^. However, stage data was only available since 2013, and missingness was still high for some cancers, which results in under-estimation of disease rates by stage and could impact trends. We therefore assessed the impact of the missing stage data through sensitivity analyses to identify cancers for which this influenced interpretation. In addition, we could not evaluate differences by tumour characteristics or subtypes because these data were not available in the public NDRS dataset used in our study. This included evaluating the impact of excluding NET or appendiceal tumours and differences by tumour location on colorectal cancer trends, and differences in trends by breast cancer oestrogen-receptor status.

There are several research and public health implications from our findings. The increase in colorectal and endometrial cancer mortality in younger adults is of particular concern. Colorectal cancer is now the second most common cause of cancer death in younger adults in England. Despite increases in incidence and mortality for endometrial cancer, fortunately, due to very good survival rates the number of deaths in younger adults is still very small (23 deaths in 2023). Although it the decreases in breast cancer mortality in younger women is very encouraging, the incidence rates of late-stage disease have increased, and breast cancer remains by far the most common cancer, and the leading cause of cancer death, in younger adults in England in 2023. As breast cancer affects primarily women, it contributes substantially to the disproportionate burden of cancer in younger women compared to men. Further investigation into the contributing factors for the increasing incidence would be important to mitigate potential future rises in mortality.

In conclusion, these analyses of incidence, mortality and stage data highlight several important public health and research priorities for cancer in younger adults.

## Data Availability

All data are available online at https://digital.nhs.uk/ndrs/data/data-outputs/cancer-data-hub/cancer-registration-statistics

## Acknowledgements

This work uses data provided by patients and collected by the NHS as part of their care and support.

## Patient and public involvement

Patients or the public were not involved in the design, conduct, or reporting of this research.

