## Supplemental Tables and Figures for "Cancer trends in England in younger adults from 2001-2023: comparing incidence, mortality and stage at diagnosis"

**Appendix Table 1:** Cancer codings and definitions as used in the NDRS website*

| **Cancer site** | **ICD Code for data by age** | **NDRS Category for data by stage** |
| --- | --- | --- |
| Brain | C71 | Not applicable |
| Breast | C50 | All Breast |
| Colorectal | C18:C20 | All Bowel |
| Endometrium | C54 | Uterus - Endometrial |
| Hodgkin Lymphoma | C81 | Hodgkin Lymphoma |
| Kidney | C64 | All Kidney |
| Leukaemia | C91:95 | Not applicable |
| Melanoma | C43 | Melanoma |
| Ovary | C56 | All Ovary |
| Prostate | C61 | All Prostate |
| Testes | C62 | All testes |
| Thyroid | C73 | Thyroid |
| * Accessed from: https://nhsd-ndrs.shinyapps.io/incidence_and_mortality/ | | |

**Appendix Figure 1:** Trends in age standardised rates (ASR) for cancer incidence from 2001-2023 by age group for colon and rectum cancers in England. Points represent by observed data and lines represent modelled trends.

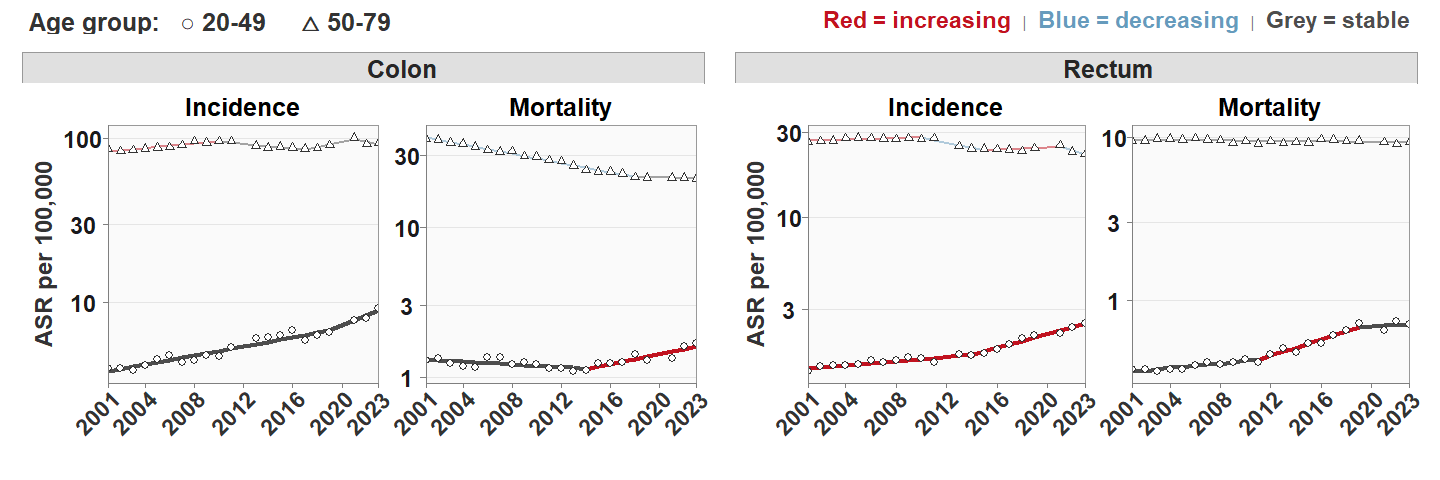

**Appendix Table 2:** Average annual percentage change (AAPC) and Annual Percentage Change (APC) for the most recent joinpoint period for 12 cancers that increased in younger adults between 2001 to 2023

| **Cancer site** | **Age** | **Incidence** | | | | **Mortality** | | | |
| --- | --- | --- | --- | --- | --- | --- | --- | --- | --- |
|  |  | **APC Years** | **APC (95% CI)** | **AAPC Years** | **AAPC (95% CI)** | **APC Years** | **APC (95% CI)** | **AAPC Years** | **AAPC (95% CI)** |
| Brain | 20-49 | 2016-2023 | -1.7 (-4.71 to 1.06) | 2001-2023 | 0.52 (0.08 to 0.95) | 2009-2023 | -0.24 (-3.8 to 3.8) | 2001-2023 | -0.73 (-1.13 to -0.32) |
|  | 50-79 | 2013-2023 | -1.04 (-2.07 to -0.48) | 2001-2023 | -0.12 (-0.46 to 0.2) | 2018-2023 | -1.2 (-6.4 to 0.15) | 2001-2023 | 0.22 (-0.01 to 0.47) |
| Breast | 20-49 | 2013-2023 | -0.32 (-1.23 to 0.17) | 2001-2023 | 0.56 (0.33 to 0.79) | 2010-2023 | -1.83 (-2.16 to -0.55) | 2001-2023 | -2.24 (-2.43 to -2.04) |
|  | 50-79 | 2003-2023 | 0.42 (-3.16 to 0.87) | 2001-2023 | 0.51 (0.33 to 0.68) | 2021-2023 | -0.46 (-1.52 to 0.56) | 2001-2023 | -2.35 (-2.44 to -2.25) |
| Colorectal | 20-49 | 2019-2023 | 6.97 (3.97 to 13.52) | 2001-2023 | 3.2 (2.79 to 3.6) | 2013-2023 | 4.11 (3.08 to 5.86) | 2001-2023 | 1.93 (1.14 to 2.74) |
|  | 50-79 | 2021-2023 | -4.36 (-7.76 to 0.57) | 2001-2023 | -0.12 (-0.5 to 0.24) | 2015-2023 | -0.94 (-1.49 to 0.75) | 2001-2023 | -1.89 (-2.1 to -1.67) |
| Endometrium | 20-49 | 2001-2023 | 2.92 (2.42 to 3.42) | 2001-2023 | 2.92 (2.42 to 3.42) | 2021-2023 | -19.86 (-39.74 to 5.33) | 2001-2023 | 3.95 (2.06 to 5.96) |
|  | 50-79 | 2011-2023 | 0.02 (-0.47 to 0.45) | 2001-2023 | 1.41 (0.86 to 1.94) | 2016-2023 | -0.29 (-2.32 to 0.82) | 2001-2023 | 1.87 (1.49 to 2.28) |
| Hodkins Lymphoma | 20-49 | 2009-2023 | 0.45 (-0.38 to 1.38) | 2001-2023 | 1.03 (0.64 to 1.39) | 2001-2023 | -6.57 (-8.66 to -4.41) | 2001-2023 | -6.57 (-8.66 to -4.41) |
|  | 50-79 | 2014-2023 | -1.22 (-2.53 to -0.25) | 2001-2023 | 1.46 (0.77 to 2.12) | 2004-2023 | -1.85 (-3.45 to -1.15) | 2001-2023 | -1.1 (-1.9 to -0.23) |
| Kidney | 20-49 | 2014-2023 | 0.98 (0.01 to 1.61) | 2001-2023 | 3.93 (3.42 to 4.44) | 2021-2023 | -11.49 (-20.97 to -1.51) | 2001-2023 | -1.69 (-2.24 to -1.1) |
|  | 50-79 | 2014-2023 | 0.82 (-0.47 to 1.61) | 2001-2023 | 2.82 (2.35 to 3.28) | 2005-2023 | -1.07 (-1.86 to -0.84) | 2001-2023 | -0.81 (-1.06 to -0.54) |
| Melanoma | 20-49 | 2013-2023 | -1.03 (-2.15 to -0.18) | 2001-2023 | 1.16 (0.54 to 1.75) | 2014-2023 | -5.76 (-7.97 to -4.38) | 2001-2023 | -2.87 (-3.55 to -2.14) |
|  | 50-79 | 2021-2023 | 7.31 (3.45 to 10.05) | 2001-2023 | 3.97 (3.38 to 4.54) | 2021-2023 | 6.42 (-1.41 to 11.36) | 2001-2023 | 0.04 (-0.67 to 0.77) |
| Ovary | 20-49 | 2001-2023 | 0.61 (0.29 to 0.92) | 2001-2023 | 0.61 (0.29 to 0.92) | 2001-2023 | -2.71 (-3.49 to -1.87) | 2001-2023 | -2.71 (-3.49 to -1.87) |
|  | 50-79 | 2001-2023 | -1.38 (-1.6 to -1.17) | 2001-2023 | -1.38 (-1.6 to -1.17) | 2001-2023 | -2.54 (-2.72 to -2.36) | 2001-2023 | -2.54 (-2.72 to -2.36) |
| Leukaemia | 20-49 | 2001-2023 | 1.11 (0.75 to 1.44) | 2001-2023 | 1.11 (0.75 to 1.44) | 2013-2023 | -1.07 (-2.76 to 7.37) | 2001-2023 | -2.62 (-3.32 to -1.88) |
|  | 50-79 | 2013-2023 | -1.61 (-2.51 to -0.88) | 2001-2023 | 0.55 (-0.04 to 1.13) | 2011-2023 | -1.91 (-5.04 to -1.4) | 2001-2023 | -1.33 (-1.57 to -1.08) |
| Prostate | 20-49 | 2011-2023 | 3.73 (-4.65 to 5.33) | 2001-2023 | 5.72 (4.74 to 6.68) | 2021-2023 | -27.66 (-50.15 to -2.04) | 2001-2023 | -4.01 (-5.47 to -2.51) |
|  | 50-79 | 2001-2023 | 1.7 (1.23 to 2.14) | 2001-2023 | 1.7 (1.23 to 2.14) | 2001-2023 | -2.03 (-2.17 to -1.88) | 2001-2023 | -2.03 (-2.17 to -1.88) |
| Testes | 20-49 | 2019-2023 | -2.91 (-12.13 to 0.21) | 2001-2023 | 0.47 (0.08 to 0.85) | 2014-2023 | 2.55 (-1.18 to 14.47) | 2001-2023 | -1.83 (-3.17 to -0.44) |
|  | 50-79 | 2001-2023 | 1.91 (1.27 to 2.53) | 2001-2023 | 1.91 (1.27 to 2.53) | 2001-2023 | 0.49 (-2.52 to 3.72) | 2001-2023 | 0.49 (-2.52 to 3.72) |
| Thyroid | 20-49 | 2013-2023 | 3.08 (2.6 to 3.42) | 2001-2023 | 5.57 (4.92 to 6.2) | 2001-2023 | 0.69 (-2.69 to 4.24) | 2001-2023 | 0.69 (-2.69 to 4.24) |
|  | 50-79 | 2015-2023 | 1.98 (-0.02 to 3.12) | 2001-2023 | 4.82 (4.28 to 5.34) | 2001-2023 | -0.15 (-0.66 to 0.39) | 2001-2023 | -0.15 (-0.66 to 0.39 |

**Appendix Figure 2:** Annual percentage change (APC) in cancer incidence in 20-49 year olds for most recent joinpoint period (in grey) for 12 cancers that increased in 20-49 year olds on average between 2001 to 2023.

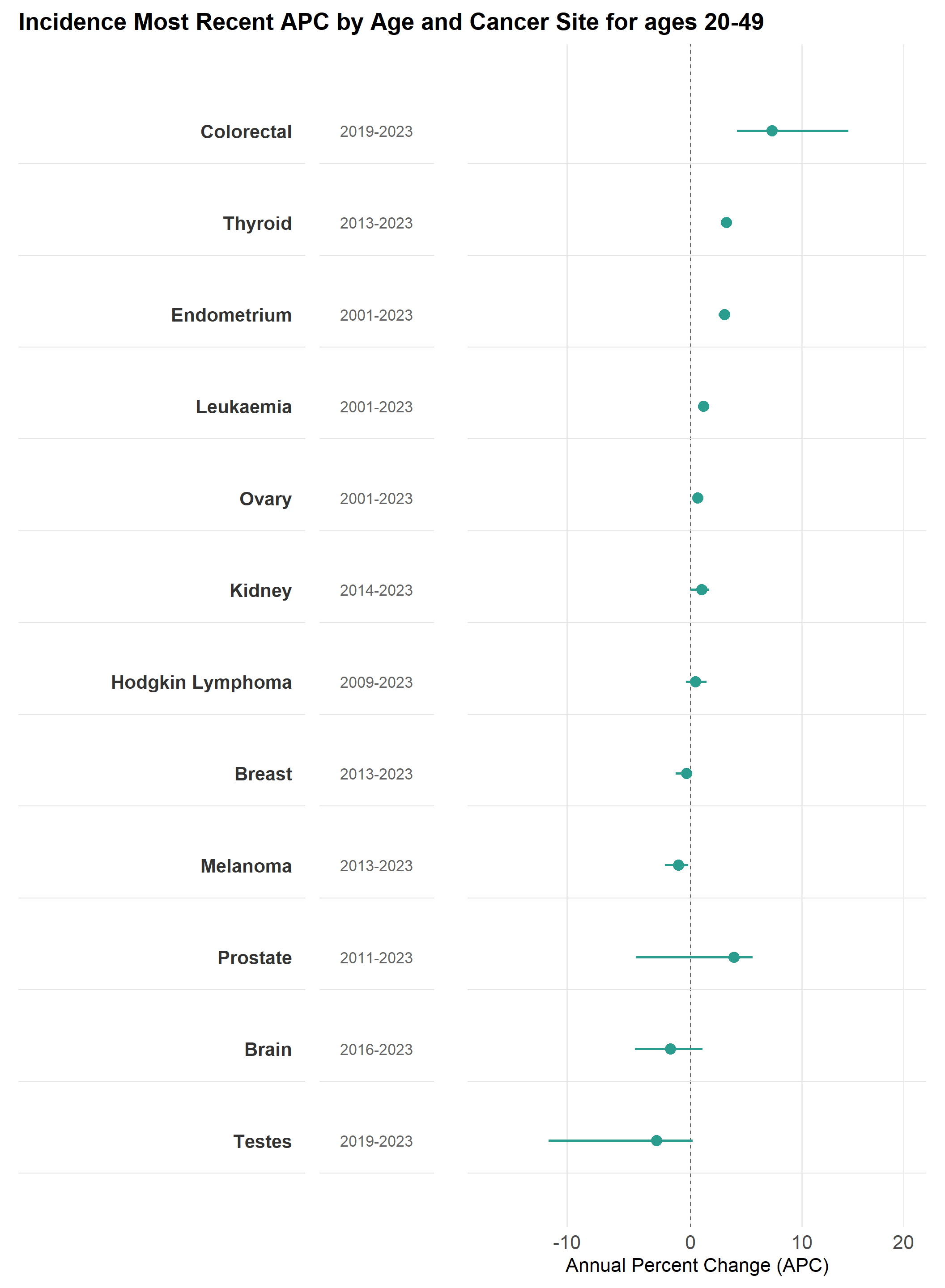

**Appendix Figure 3:** Trends in age standardised rates (ASR) for cancer incidence from 2001-2023 by ten-year age group (20-29; 30-39; 40-49; 50-59; 60-69; 70-79)in England for **(A)** cancers with AAPCs indicating increasing incidence and mortality, **(B)** cancers with AAPCs indicating increasing incidence and constant mortality, and **(C)** cancers with AAPCs indicating increasing incidence and decreasing mortality. Points represent observed data, and lines represent modelled trends.

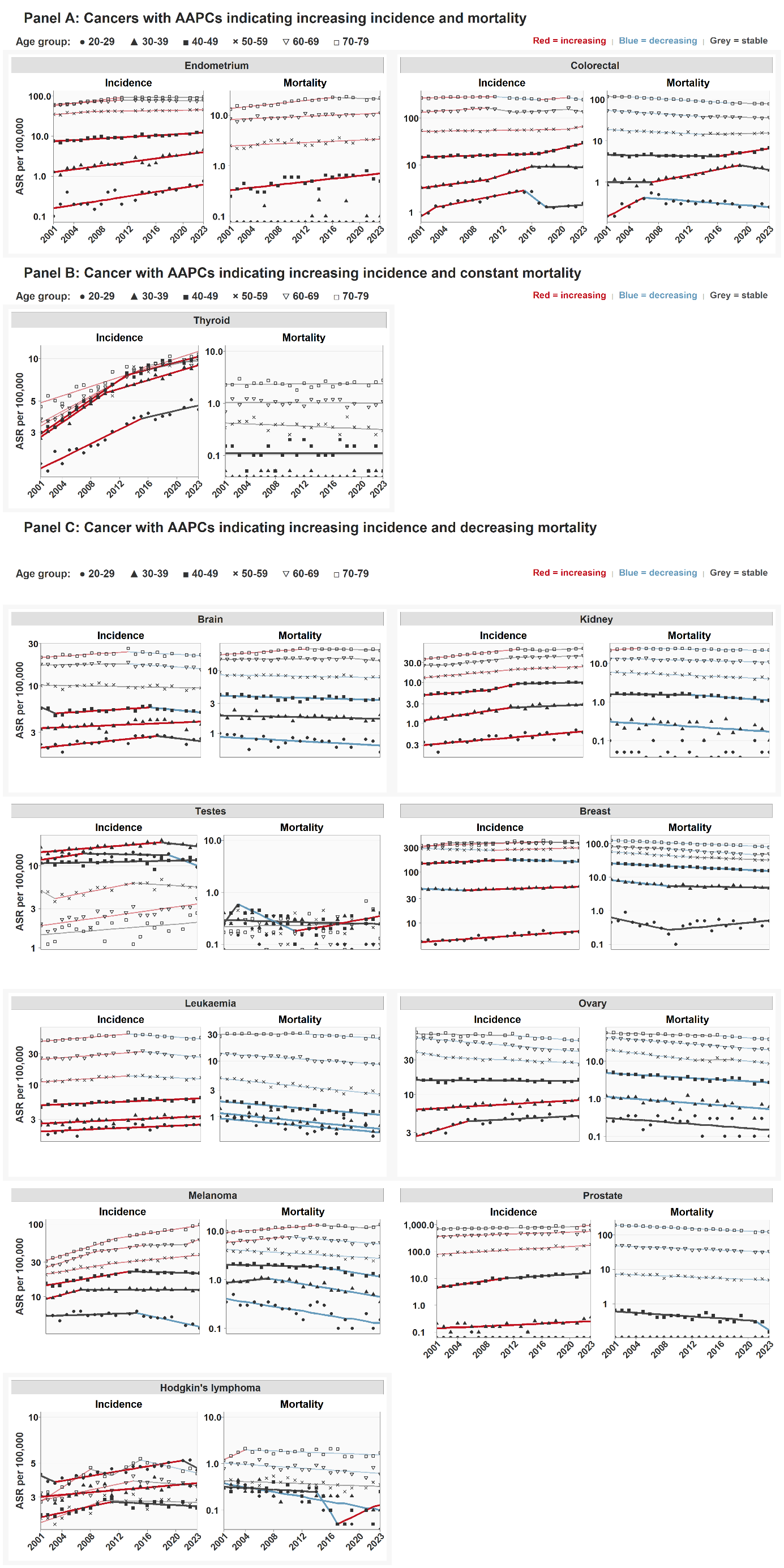

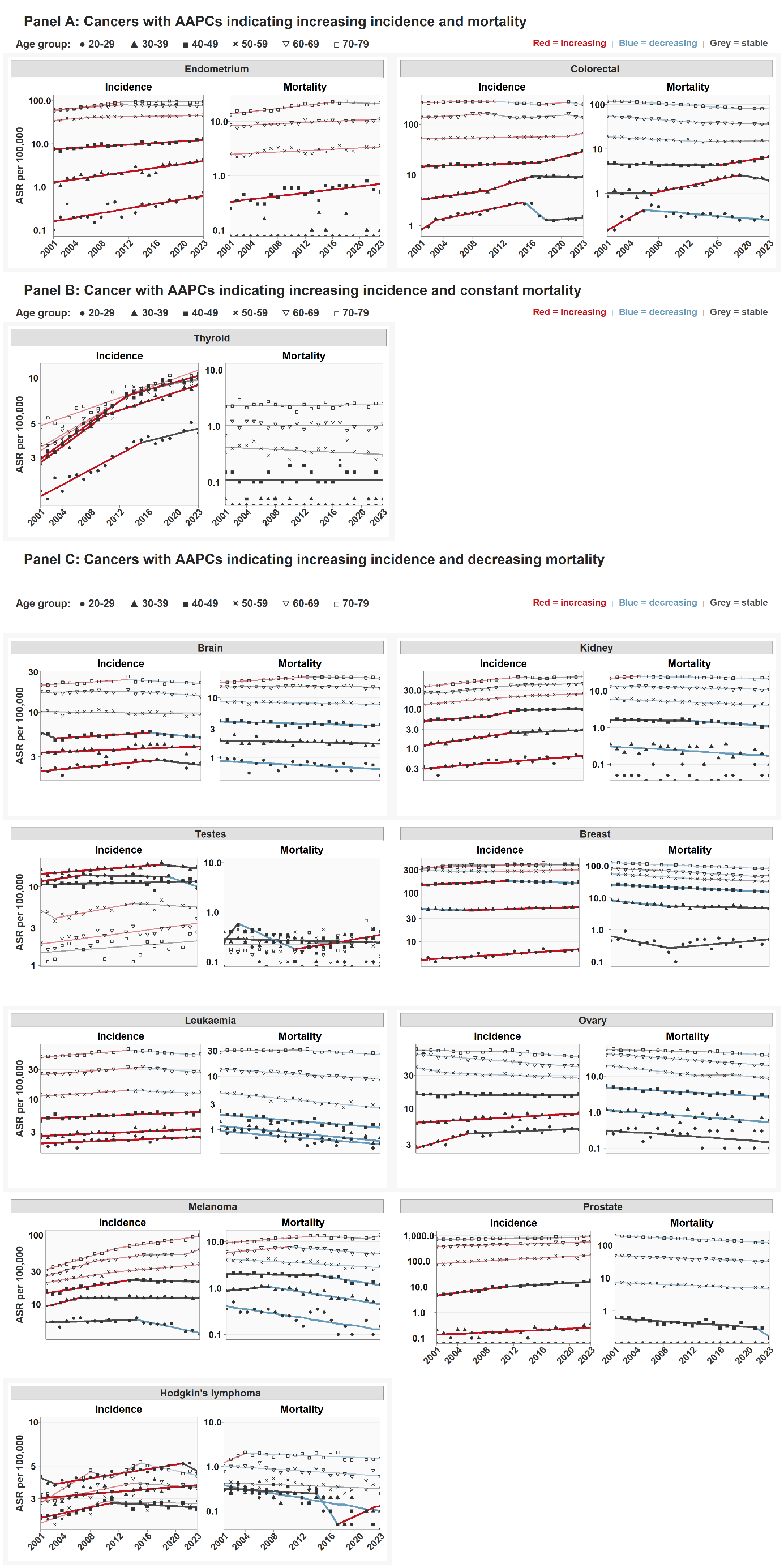

**Appendix Table 3:** Percent change (%) in age standardised incidence rates (ASR) by cancer site and stage at diagnosis for 20-49 year olds from 2013 to 2023

| **Cancer site** | **Stage** | **ASR 2013** | **ASR 2023** | **% Change (2013–2023)** |
| --- | --- | --- | --- | --- |
| Breast | Early Stage | 18.9 | 17.5 | -7.3 |
|  | Late Stage | 4.1 | 4.4 | 7.0 |
|  | Unstageable / Missing | 6.0 | 6.8 | 13.0 |
| Colorectal | Early Stage | 1.0 | 1.4 | 47.0 |
|  | Late Stage | 1.6 | 2.7 | 64.8 |
|  | Unstageable / Missing | 0.7 | 0.9 | 19.7 |
| Endometrium | Early Stage | 1.0 | 1.3 | 27.1 |
|  | Late Stage | 0.1 | 0.3 | 78.9 |
|  | Unstageable / Missing | 0.3 | 0.6 | 88.2 |
| Hodgkin Lymphoma | Early Stage | 0.5 | 0.5 | -0.1 |
|  | Late Stage | 0.4 | 0.5 | 22.1 |
|  | Unstageable / Missing | 0.3 | 0.2 | -30.0 |
| Kidney | Early Stage | 0.7 | 0.8 | 19.2 |
|  | Late Stage | 0.3 | 0.3 | 2.3 |
|  | Unstageable / Missing | 0.5 | 0.6 | 24.3 |
| Melanoma | Early Stage | 3.8 | 3.3 | -12.2 |
|  | Late Stage | 0.3 | 0.4 | 29.8 |
|  | Unstageable / Missing | 0.9 | 0.9 | 4.0 |
| Ovary | Early Stage | 1.8 | 1.6 | -8.5 |
|  | Late Stage | 0.8 | 0.9 | 6.1 |
|  | Unstageable / Missing | 1.6 | 1.9 | 12.9 |
| Prostate | Early Stage | 0.8 | 1.4 | 86.7 |
|  | Late Stage | 0.3 | 0.4 | 24.5 |
|  | Unstageable / Missing | 0.3 | 0.6 | 94.3 |
| Testes | Early Stage | 2.7 | 3.6 | 34.6 |
|  | Late Stage | 0.2 | 0.3 | 22.7 |
|  | Unstageable / Missing | 3.0 | 0.8 | -73.6 |
| Thyroid | Early Stage | 0.8 | 1.7 | 106.1 |
|  | Late Stage | 0.1 | 0.0 | -79.3 |
|  | Unstageable / Missing | 1.6 | 1.5 | -4.9 |

**Appendix Table 4:** Percentage (%) of cancers with missing stage at diagnosis by year of diagnosis, cancer and age at diagnosis: ordered by % of missing data in 20-49 year olds.

| **Cancer site** | **Age at diagnosis (yrs)** | **2013** | **2015** | **2018** | **2021** | **2023** |
| --- | --- | --- | --- | --- | --- | --- |
| Thyroid | 20-49 | 55 | 31 | 23 | 33 | 40 |
|  | 50-79 | 54 | 31 | 24 | 34 | 37 |
| Ovary | 20-49 | 30 | 18 | 19 | 30 | 31 |
|  | 50-79 | 31 | 18 | 17 | 32 | 34 |
| Kidney | 20-49 | 31 | 15 | 17 | 27 | 30 |
|  | 50-79 | 26 | 11 | 13 | 28 | 29 |
| Prostate | 20-49 | 21 | 12 | 11 | 21 | 18 |
|  | 50-79 | 17 | 9 | 8 | 19 | 19 |
| Breast | 20-49 | 17 | 8 | 11 | 16 | 17 |
|  | 50-79 | 15 | 7 | 9 | 17 | 17 |
| Colorectal | 20-49 | 19 | 11 | 11 | 16 | 17 |
|  | 50-79 | 17 | 10 | 8 | 12 | 13 |
| Hodgkin Lymphoma | 20-49 | 22 | 8 | 10 | 19 | 16 |
|  | 50-79 | 19 | 5 | 8 | 18 | 13 |
| Endometrium | 20-49 | 15 | 7 | 8 | 17 | 15 |
|  | 50-79 | 17 | 7 | 10 | 23 | 22 |
| Melanoma | 20-49 | 17 | 10 | 9 | 14 | 15 |
|  | 50-79 | 13 | 6 | 6 | 12 | 15 |
| Testes | 20-49 | 43 | 13 | 12 | 15 | 13 |
|  | 50-79 | 41 | 11 | 11 | 14 | 12 |

**Appendix Figure 4:** Sensitivity analysis for age standardised rates for cancer incidence in 20-49 year olds in 2013 and 2023 by stage at diagnosis by cancer in 3 scenarios **(1)** missing stage and unstageable cancers included as distinct category **(2)** missing stage and unstageable cancers coded as “Early Stage” disease **(3)** missing stage and unstageable cancers coded as “Late Stage” disease

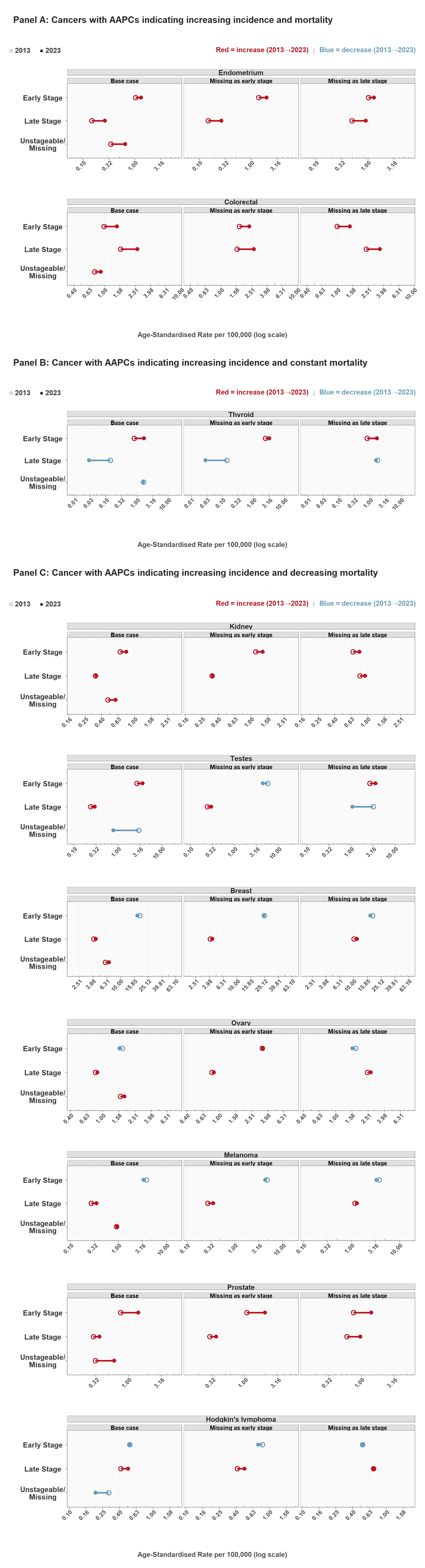

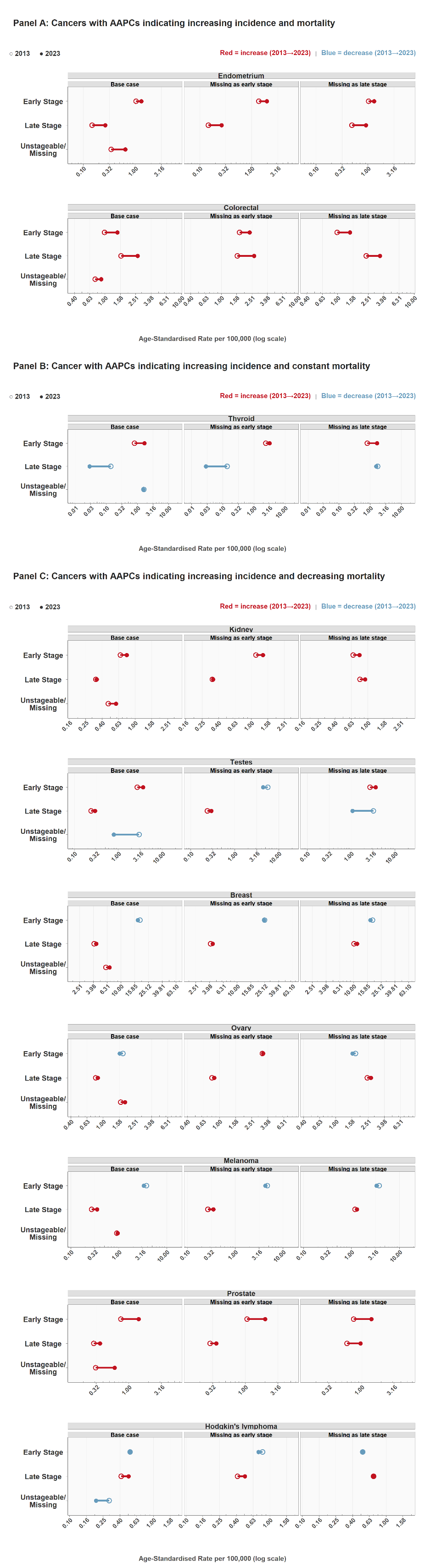
